# Does The Reformed Cancer Drug Fund Generate Evidence On Effectiveness? A Cross-sectional Analysis On Publicly Accessible Documentation

**DOI:** 10.1101/2020.03.06.19014944

**Authors:** Helen Macdonald, Ben Goldacre

## Abstract

**Introduction:** The Cancer Drugs Fund (CDF) was reformed in 2016 with an ambition to generate new evidence on effectiveness, and to review existing drugs in the fund. We set out to evaluate: whether drugs transitioning from the old CDF were re-reviewed as planned; whether new drugs have a “data collection arrangement” (DCA) as planned; and whether evidence generated under the DCA using routine data from the “Systemic Anti-Cancer Treatment” (SACT) database was of high quality.

**Methods:** We accessed documents from NHS England, Public Health England and NICE at August 2018. We calculated the proportion of old CDF drugs re-reviewed, and of new drugs and indications with a DCA. We described key features of the DCAs. For all SACT studies we set out to obtain a protocol in order to analyse the quality of the planned methods.

**Results:** 47 old drugs and indications transitioned to the new CDF. For 14 there was no evidence of a re-review; 9 of these remain under CDF at August 2019 (all off-label uses). 33 had marketing authorisations: 22 of these had a review completed by September 2017 as planned (67%). 20 new drugs and indications entered the CDF by August 2018: 19 had a DCA; one (off-label) had no DCA or equivalent. All DCAs identify uncertainty about overall survival; all express an intent to conduct observational analysis using SACT data; SACT data was central to decision-making for 6 (32%). We were able to find 0 protocols of the 19 planned SACT studies (0). Following Freedom of Information requests we were told these protocols are prepared after the data are collected, and posted with the reappraisal: however we could not locate any protocol for either of the two published re-appraisals. We were therefore unable to assess the quality of the methods in any of the proposed SACT studies.

**Conclusions:** The revised CDF has not been implemented as planned. Reporting of observational analyses in SACT data fall substantially short of best practice, and the full methods used cannot be established. There is very little information in the public domain around evaluation of off-label uses. Lastly, SACT data itself does not appear to be able to support clinical decision-making in the manner suggested by the CDF policy documents. NHS England should review the conduct of the fund, but also the planning, as unrealistic commitments may have been made.

## Introduction

The Cancer Drugs Fund (CDF) was launched in 2010 and added complexity to the process whereby new high-cost medicines are subject to a single technology appraisal. NICE appraisal assesses their effectiveness, cost-effectiveness, and value to the UK National Health Service and drugs are either recommended or declined for use on the NHS. The aim the CDF was to accelerate access for new high cost treatments and thereby improve outcomes for cancer patients (National Audit Office, 2015), on the grounds that society might view cancer as a disease worthy of special treatment (Linely and Hughes, 2012). By 2014 approximately 1 in 5 people starting chemotherapy took a CDF drug (National Audit Office, 2015). Treatments on CDF bypass the usual decision-making processes: some were awaiting NICE assessment, others had been appraised and actively declined (The House of Commons Committee of Public Accounts, 2016) (National Audit Office, 2015).

The fund has attracted extensive criticism because impact proved difficult to quantify; costs grew rapidly when drugs did not leave the fund as planned; the processes were opaque (Dixon et al 2016); and other analyses of pivotal trials for the included drugs showed that many fell short of clinically important thresholds (Aggarwal *et al*., 2017). Criticism culminated in two major national investigations that questioned the CDF’s premise, processes, sustainability and overall value (The House of Commons Committee of Public Accounts, 2016) (National Audit Office, 2015). This triggered a consultation that led to reform of the CDF (NICE, 2015).

Under the new CDF there was a to be a stronger focus on transparency, process and value for the public, with the process reframed as a “managed access fund” generating new evidence. Two key documents from NICE and NHS England outline broadly how the CDF is intended to work (NICE, 2016f and NHS England Cancer Drugs Fund Team, 2016). NICE has resumed control of appraising licensed drugs and indications, delivering expedited assessments; while NHS England has created a parallel process for “off label” uses of treatments outside their current marketing authorisation (commonly for use in children). Drugs enter the reformed CDF if there is too much uncertainty to reach a clear decision about them (NICE, 2016g). New evidence to resolve uncertainty about the drug must be generated within two years of the drug entering the fund. Once these data are available, the drug undergoes a second appraisal (NICE, 2016e).

The additional evidence can come from ongoing trials, or be generated through a new system conducting observational analyses of “real-world data” collected in the Systemic Anti-Cancer Treatment database (SACT) and linked to other registries such as “hospital episode statistics” and deaths (National Cancer Registration and Analysis Service, 2015). Public Health England are responsible for delivering SACT studies, which may address uncertainties around efficacy, adverse events, tolerability and optimal dosing. Plans for resolving outstanding uncertainties must be described in a Data Collection Agreement (DCA) when a treatment enters the CDF. In addition to this new process for emerging treatments and indications, it was announced that all drugs transitioning from the old CDF would be re-reviewed by NHS England and NICE before September 2017.

The NHS model of technology appraisal by NICE and national price negotiations has been internationally influential (Edwards et al, 2019): the CDF is likely to attract similar interest and perhaps influence. Many countries face challenges around accelerated access to innovative treatments; and around rapidly evaluating new treatments in a challenging landscape with escalating costs, competing patient interests and complex arguments around what constitutes good quality evidence for benefit or harm. We therefore set out to assess whether the new CDF was being implemented as planned for old drugs transitioning into the new system (NICE, 2016b, appendix 2); and for current CDF drugs (NHS England, 2018b, appendix 1); and to assess the extent and quality of the new “real world evidence” generated by the NHS through the SACT analyses.

## Methods

### Re-reviewing old CDF drugs by September 2017

The NICE website contains a list of all old drugs transitioning into the new CDF system, and their current appraisal status (NICE, 2016b). For each drug and indication in this list we searched for publicly available decision and supporting documentation using the NICE and NHS England websites, and Google, in order to assess compliance with the new CDF policy. For licensed indications of approved treatments, the CDF documents state: “All drugs on the previous CDF as of 31 March 2016 will be reconsidered or appraised by NICE over the course of the next 18 months”. Our compliance standard for this was the presence of a NICE technology appraisal (or equivalent for off-label drugs) by September 2017. For unlicensed indications or treatments, the commitments are less clear, stating: “NHS England will be responsible for overseeing a process for considering the commissioning of cancer drugs for off-label indication use… NHS England is in the process of commissioning clinical evidence reviews to inform their consideration… this will be a similar procedure to that used by NICE” (NHS England Cancer Drugs Fund Team, 2016). Our compliance standard here was the presence of health technology appraisal decision and supporting documents on the NHS England or NICE website.

### Presence of Data Collection Agreements for new CDF treatments

All treatments and indications funded by the CDF are required to have a “Data Collection Agreement” (DCA) document that describes what new evidence will be generated to resolve outstanding uncertainties (NHS England Cancer Drugs Fund Team, 2016). We downloaded the list of all new treatments and indications approved under the CDF from NHS England on 31/8/18 ((NHS England, 2018b, Supplementary Material). For each drug and indication we searched for publicly available documentation using the NICE and NHS England websites, and on Google, to assess for the presence or absence of a DCA.

### Descriptions of Clinical Uncertainty in DCAs

DCAs are required to describe the uncertainties they are setting out to resolve. The CDF policy documents contain very little information on how this should be expressed or categorised, and the contents of the DCAs themselves are typically free text. Therefore we used Morrell *et al’s* classification of uncertainties in cancer drug technology appraisals (Morrell *et al*., 2018) to provide a clearer structure. For each indication we read the DCA and recorded whether the outstanding uncertainties were caused by: immature survival data; lack of relevant comparators; problems with design of prior trials; problems with external validity of prior trials; shortage of quality of life data; reliance on observational data; lack of evidence on optimum treatment duration; lack of evidence on adverse events.

### Availability of protocols or detailed methods for SACT analyses

For each treatment we noted whether the primary source of evidence in the Data Collection Agreement was a trial, an SACT study, or “other” study. The CDF scheme emphasises the importance of SACT data and states that these observational analyses will “always” occur, may be supplemented by trial data, but could be the only source of data for future decision making (NICE, 2016f and NHS England Cancer Drugs Fund Team, 2016). We aimed to investigate the transparency and methodological standards around the new system for “real-world” observational data from SACT further. Our criteria for assessing this was that there should be a pre-specified protocol for the analysis of the SACT data (World Medical Association, 2013), either in the DCA, or clearly signposted from it (for example from beneath the DCA headings “outcome data” and “data analysis plan”).

### Availability and quality of SACT studies to resolve uncertainties

We set out to collect all available documentation describing the planned data analysis by PHE for critical appraisal by research methodologists, and to extract themes around rigour and appropriateness of the methods proposed for the specific clinical questions posed. However, as described below, no detailed documentation could be found for any of the “real world evidence” analyses planned.

## Results

### Reviewing old CDF drugs by September 2017

47 treatments and indications transitioned from the old CDF into the new scheme and should have had a final decision by September 2017 (NICE, 2016b and appendix). Figure 1 shows the ultimate outcomes for these treatments. 14 of the 47 drugs and indications had no marketing approval from the European Medicines Agency and should be covered by NHS England’s “evidence review” process for assessing “off label” indications in the CDF. We could find no clear evidence of assessment by NHS England for any of these 14 indications in the public domain: neither plans for these, nor the results. Box 1 narrates elements of four publicly available documents published by NHS England after the September 2017 deadline. It is unclear to us whether these are reviews of four transition drug and indication pairs. At August 31st nine of these 14 indications were still listed as being reimbursed on the CDF list; five were no longer in the fund.

**Figure 1.**
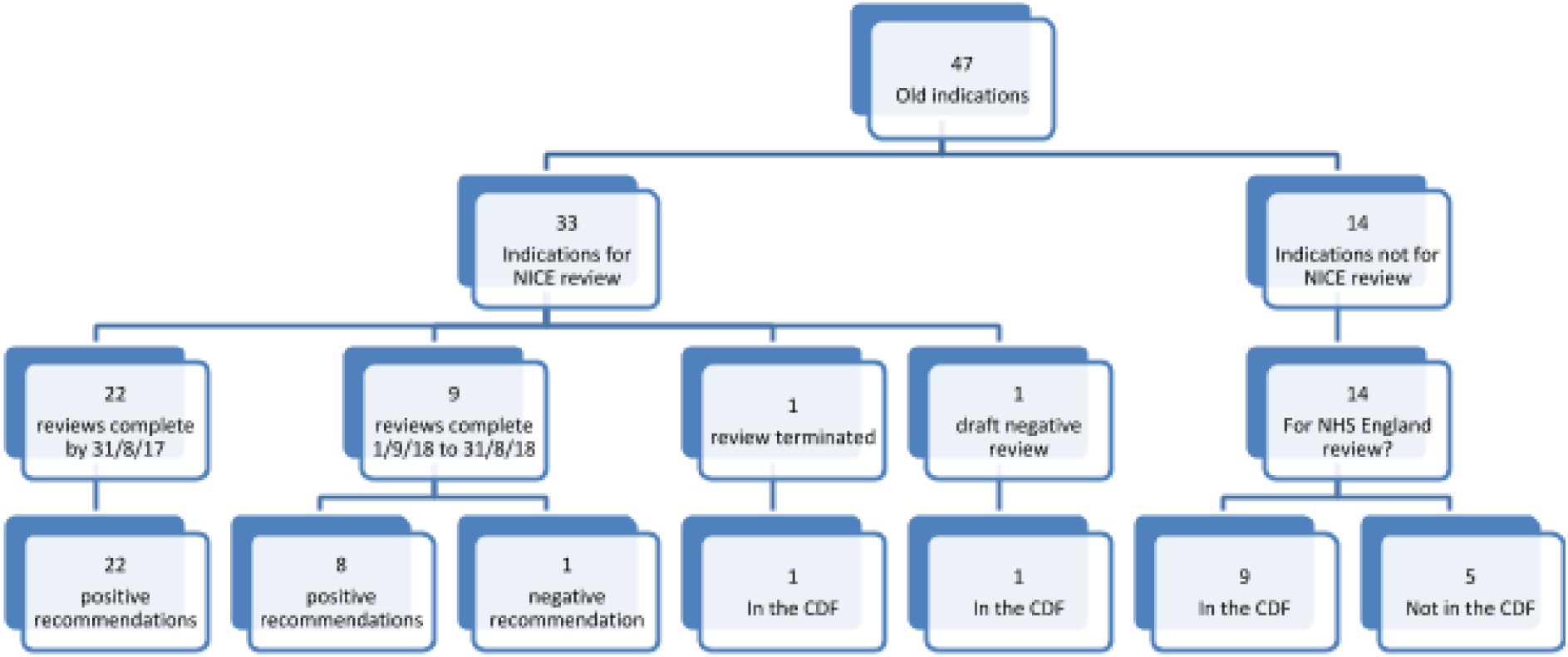
Outcomes (if known) for old CDF indications that entered the reformed fund by 31/8/18 Footnote to figure: Indications can only be reviewed by NICE if they have a licence for that indication. Those without a license for that indication were to be reviewed by NHS England.

#### Box 1

Four publicly available documents which may represent reviews of two transition CDF drugs (bendamustine and clofarabine)

Each of the four “Clinical Policy Review” documents are publicly available on the NHS England’s website in the publication section.

There are three documents including the keyword bendamustine in the title and published on the NHS England site which may be relevant:

- None of the three documents is clearly labelled as a review of a transition CDF drug, however each lists the CDF site and the current list in “documents which have informed this policy”
- Each of the documents is about bendamustine with rituximab, rather than bendamustine alone, which is the drug on transition list
- The indications on the list and in the documents are an imprecise match (CDF list states: lymphoma first line, lymphoma second line and Non-Hodgkin lymphoma. NHS England documents list: relapsed and refractory non-mantle cell lymphoma (all ages); first line treatment of mantle cell lymphoma (all ages) and first line treatment of advanced indolent non-Hodgkin’s lymphoma (all ages).)
- There is a fourth indication for bendamustine (alone) on the CDF transition sheet and there is not a matching clinical commissioning policy document for this
- These documents were published after the review deadline in July 2018.

There is one document including the drug clofarabine as a keyword on NHS England’s site

- This document is not clearly labelled as a review of a transition CDF drug and was published in November 2018
- This document does not mention other CDF resources such as the list or website, as the examples above did
- The indication in the document reads “refractory or relapsed acute myeloid leukaemia (AML) as a bridge to stem cell transplantation (all ages))”. On the transition document it reads “leukaemia (acute myeloblastic) second line”.
- In the section on epidemiology it says 48 people received the drug through the cancer drugs fund in 2017, suggesting that the drug had at least one indication in the CDF then. It had two indications on the CDF list of August 2018.
- This document was published after the review deadline in November 2018.

Documents:

Clinical Commissioning Policy: Bendamustine with rituximab for first line treatment of mantle cell lymphoma (all ages). Available: https://www.england.nhs.uk/wp-content/uploads/2018/07/1630-bendamustine-and-rituximab-for-mcl.pdf (accessed 3/12/19

NHS England (2018) Clinical Commissioning Policy: Bendamustine with rituximab for relapsed and refractory mantle cell lymphoma (all ages). Available: https://www.england.nhs.uk/wp-content/uploads/2018/07/1604-bendamustine-with-ritxumab-for-mcl.pdf (accessed 3/12/19)

Clinical commissioning policy: Bendamustine with rituximab for first line treatment of advanced indolent non-Hodgkin’s lymphoma (all ages). Available: https://www.england.nhs.uk/wp-content/uploads/2018/07/1605-bendamustine-with-rituximab-for-nhl.pdf (accessed 3/12/19)

NHS England (2018) Clinical Commissioning Policy: Clofarabine for refractory or relapsed acute myeloid leukaemia (AML) as a bridge to stem cell transplantation (all ages). Available: https://www.england.nhs.uk/wp-content/uploads/2018/11/clofarabine-refractory-relapsed-acute-myeloid-leukaemia.pdf (accessed 3/12/19)

33 of the 47 drugs and indications had marketing authorisations from the European Medicines Agency and were therefore eligible for assessment by NICE: of these 33 had a planned review, 22 had completed by September 2017 as required (67%); 31 had completed by September 2018 (94%); one had a draft negative recommendation; and one was terminated as the company submitted no data.

### Presence of Data Collection Agreements for new CDF treatments

20 new drugs had entered and remained in the reformed CDF by 31st August 2018. All should have a DCA. 19 treatments had a marketing authorisation for the indication covered under CDF, and therefore should have had a DCA created by NICE: all 19 were compliant on this requirement. One drug which was approved for off-label use under CDF should have had the equivalent of a DCA from NHS England’s parallel review process for off-label drugs: we were unable to locate such a document.

### Descriptions of Clinical Uncertainty in DCAs

The 19 DCAs described a wide range of clinical uncertainties for the treatments covered. We used relevant elements of Morrell’s classification system to characterise the uncertainties described in the documents (Appendix 3) (Morrell *et al*., 2018). 19 DCAs (100%) described outstanding uncertainties due to immature survival data; 9 (47%) described uncertainties due to lack of an appropriate comparator; 9 (47%) described uncertainties due to lack of relevant patient population; 7 (37%) described uncertainty around duration of use of the treatment; 3 (16%) described uncertainty around quality of life. All DCAs outlined plans to use SACT data; however, observational analysis of SACT data was the primary source of information to guide decision making in only 6 CDF approvals (32%).

### Availability of protocols or detailed methods for SACT analyses

We evaluated all 19 DCAs planning to use SACT data. We were unable to locate any protocol, analysis plan, or any other document that substantially described the planned analyses for the SACT data, for any of the 19 DCAs. We therefore sent Freedom of Information (FOI) Act requests to ask NHS England, PHE and NICE for the protocol for all planned SACT studies. The aim of this correspondence was to establish whether any such protocols had been created; to establish whether protocols for observational analyses had been devised prior to data collection and analysis, as per best practice; and ideally to read and critically appraise the protocols for the planned analyses on SACT data. These FOI requests are registered on “What do they know” (2015). From this correspondence (appendix 4), we were able to establish that protocols do or will exist but will not be shared before the second appraisal of the drug. We were told that the methodology of the study will be written after data collection, but before analysis; and that the methodology or protocol will be publicly available with the technology appraisal on the NICE website, when the evaluation is published (but not before).

### Availability and quality of SACT studies to resolve uncertainties

As stated above, no protocols were available for any observational analyses planned in SACT for any of the CDF drugs and indications. We were able to identify two examples of completed re-appraisals by NICE, where according to PHE’s FOI response there should have been a protocol for the SACT analysis published after the analysis was conducted and completed: pembrolizumab for non-small-cell lung cancer (TA531) (NICE, 2018d); and brentuximab vedotin for CD30-positive Hodgkin lymphoma (TA524) (NICE, 2018b). For both drugs we were unable to locate any such protocol on NICE’s website, with the HTA report, or in the linked documentation. The pembrolizumab DCA of 31/05/2017 describes a planned SACT analysis, but the NICE re-appraisal for this treatment 18/07/18, where the SACT analysis should be reported alongside its protocol, does not mention SACT data at all: this SACT analysis would therefore appear to be a case of non-publication, or publication bias. The Brentuximab DCA 28/04/2017 describes a planned SACT analysis; the NICE re-appraisal of this treatment 13/06/2018 does not describe the SACT analysis, but does briefly describe a “real-world UK” observational study including 78 people; however we were unable to locate a protocol or description of methods used for this analysis in the NICE documents, or linked papers.

We updated the search for re-reviews published of new CDF drugs up until 29/11/19. No further re-reviews and linked technology appraisals had been published by NICE.

### Additional observations

Throughout the process of conducting this research we were struck by the inaccessibility of information around the CDF. Documents were hard to locate. There is no one stop website where all of the information is collected. The plans and operating models for the CDF were dispersed across a large number of long partially overlapping documents, and we were unable to identify a single succinct description of how the CDF should operate that showed both off-label drugs and those with a license for that indication. The challenges in locating information on evaluation of individual treatments were compounded by the fact that information apparently intended to be public (such as the methods of the SACT studies) was commonly not available. Furthermore in 6 (32%) of DCAs there were two or more areas which had been redacted (see appendix 5): these redactions appeared to be inconsistent, but we cannot know if further important information relevant to our analysis was withheld by this process of redaction.

## Discussion

### Summary

The revised CDF has not been implemented as planned. Re-review and public display of decisions of old drugs did not occur on time, particularly for off-label drugs. DCAs were produced but contained little or no information about the observational analyses planned in SACT. No protocols were available for any SACT analyses. We were told these were only intended to be shared after the CDF review has completed; however for the two treatments where the CDF review has completed, neither the methods nor the results of the relevant SACT analyses appear to have been disclosed. Documents were hard to locate, lacked succinct descriptions of key CDF processes, and were commonly redacted.

### Strengths and weaknesses

To our knowledge this is the first evaluation of the new CDF. We are independent of the scheme’s conception and delivery. A possible weakness is that we were unable to obtain all documents: however we regard the inaccessibility of the relevant CDF documents as a key finding of the research. Another possible weakness is that we may have misunderstood or overlooked some elements of the operational plans for the CDF: again, we view the discursive and relatively inaccessible documentation of these plans as a key finding of our research. A final possible weakness is that we used the CDF list of August 2018. However this is beyond the date by which the relevant components of the CDF should have been implemented; and the responses to our FOI requests clearly show that the inaccessibility of SACT analyses will not have changed subsequent to these dates, as this is a matter of policy rather than delayed implementation. We searched for additional evidence of posting protocols and SACT data from 31/08/18 to 29/11/19 with repeat technology appraisals, however we were unable to find evidence that further appraisals had been published, although several are due in the coming months.

### Comparison to other work and remaining uncertainties

We are aware of no other substantial work evaluating the implementation of the new CDF. An internal review by the organisers of the CDF scheme itself has been conducted and is due to report in 2018/19 (see appendix 5); as of 29/11/19 we can find no evidence of this review being published. The process of the fund was only one of a number of problems with it. Recent international research suggests that the premise of the fund also remains uncertain. A systematic review of studies about public opinion of the value of treating conditions at the end of life (Shah *et al*., 2018) suggests that people are divided on whether they should be singled out for special treatment. Of 23 suitable studies 11 studies suggested that there was not opinion to support an end-of-life premium (48%), eight studies suggested there was (35%), and 4 were inconclusive (17%). Another systematic review found that people viewed cancer as a ‘special’ condition; but when presented with the opportunity cost of that choice this does not necessarily translate into an increased willingness to pay for care (Morrell *et al*., 2017).

### Policy implications

In our view the ambitions of the revised CDF were questionable, in particular the plan to address questions of effectiveness using observational data. Such analyses are subject to a substantial risk of bias due to the many discretionary analytic choices, the possibility of conscious or unconscious bias in analysis design, the risk of selective reporting, and the risk of “p-hacking”. Best practice is to ensure prespecification of methods, and publication of a full protocol. The risks around *post hoc* data analysis are exacerbated where the objective is to detect modest benefits such as those from CDF treatments, because these weak signals are more liable to be overwhelmed by bias, confounding, or statistical noise. In this light, while we were concerned to find no protocols for any planned analyses, we were particularly concerned by the apparent non-publication of one, if not two, completed SACT analyses. Those involved in SACT database analyses have recently published a paper profiling the database (Bright et al, 2019). From this we also note that problems with the completeness and accuracy of the underlying SACT data itself may further undermine the feasibility of the revised CDF plans.

We identify two further areas of concern. Firstly, we can see no justification for treating off-label indications differently, and specifically no justification for holding such treatments to lower standards around transparency of process, and publication of HTA outcomes. These off-label uses commonly include drugs used in the care of children with cancer. Secondly, we are very concerned by the relative inaccessibility of information on the processes and specific outcomes of the CDF system.

In our view the CDF should be reviewed. All SACT protocols and analyses should be shared in full, immediately, and clearly linked to (or published in) the HTA documents to maximise discoverability. More broadly, in our view the inclusion of observational analyses itself seems to have been poorly thought through: we strongly recommend that NHS England or other independent parties review the processes leading up to these plans, to assess whether the failings were foreseeable.

### Conclusion

The revised CDF has not been implemented as planned, and there are signs of a troubling lack of transparency. NHS England should review the conduct of the fund, but also the planning, to assess whether unrealistic commitments around observational analysis of data were made.

## Data Availability

The data are attached to the main paper.

## Acknowledgements

This project formed part of HM’s MSc course, the fees for which were 50% funded by The BMJ.

## Ethical approval

This study uses exclusively open, publicly available data, therefore no ethical approval was required.

## Contributorship

BG conceived the study. HM BG designed the methods. HM collected and analysed the data with methodological and interpretation input from BG. HM BG drafted the manuscript. BG supervised the project. HM and BG are guarantor.

## Patient and public partnership

Patients were not involved in the design, or conduct, or reporting of our research. We plan to disseminate findings to relevant patient and public groups.

